# ^18^F-FDG PET/CT metabolic parameters predict prognosis in pancreatic ductal adenocarcinoma after neoadjuvant chemotherapy

**DOI:** 10.64898/2026.02.28.26347307

**Authors:** Linghe Zhang, Luqiang Jin

## Abstract

This study aimed to evaluate the prognostic value of quantitative analysis of ¹⁸F-FDG positron emission tomography (PET)/computed tomography (CT) metabolic parameters in patients with pancreatic ductal adenocarcinoma (PDAC) after neoadjuvant chemotherapy (NACT). A retrospective analysis was conducted on the clinical and imaging data of 44 patients with pathologically confirmed PDAC who received NACT. All patients completed standard chemotherapy regimens and underwent ¹⁸F-FDG PET/CT examinations within 2 weeks before and after chemotherapy. Multiple metabolic parameters of lesions were extracted, their percentage changes were calculated, and the optimal cut-off values for each parameter were determined. Kaplan–Meier survival analysis and Cox proportional hazards regression analysis were applied to explore the prognostic value of the metabolic parameters, and the prognostic stratification performance of PET Response Criteria in Solid Tumors (PERCIST) 1.0 was compared with that of Response Evaluation Criteria in Solid Tumors (RECIST) 1.1. PERCIST 1.0 demonstrated significantly superior prognostic stratification compared with RECIST 1.1. A peak standardized uptake value corrected for lean body mass (SULpeak_2_) > 3.07 and a percentage change in SULpeak between pre- and post-treatment scans (ΔSULpeak%) ≤ 37.66% were identified as independent risk factors for poor prognosis. Furthermore, SUL-related parameters exhibited markedly better predictive efficacy than traditional metabolic parameters such as the standardized uptake value and metabolic tumor volume. Quantitative analysis of ¹⁸F-FDG PET/CT metabolic parameters can effectively predict prognosis in PDAC after NACT, and PERCIST 1.0 is a more optimal criterion for efficacy and prognostic assessment. A post-NACT SULpeak > 3.07 and ΔSULpeak% ≤ 37.66% were core independent indicators for predicting poor prognosis in these patients.

## 1. Introduction

Pancreatic ductal adenocarcinoma (PDAC) is the most common type of pancreatic cancer, characterized by extremely high malignancy and poor prognoses. Currently, the 5-year survival rate of pancreatic cancer is less than 8%[1, 2]. Although radical surgical resection can prolong patients’ survival, nearly 80% of patients present with unresectable disease at the time of diagnosis. Neoadjuvant chemotherapy (NACT) not only improves the R0 resection rate in patients with borderline resectable pancreatic cancer but also enables some patients with locally advanced pancreatic cancer to receive radical surgical treatment[3–6]. The Response Evaluation Criteria in Solid Tumors version 1.1 (RECIST 1.1), based on computed tomography (CT) and magnetic resonance imaging (MRI), provides a valuable reference for current efficacy and prognostic assessment[7, 8]. However, several studies have reported that treatment responses evaluated by RECIST 1.1 are not associated with patients’ overall survival (*OS*)[9]. This may be attributed to the unique tumor microenvironment of PDAC, as well as tumor stromal fibrosis and inflammatory edema of adjacent tissues induced by chemotherapy[10]. These factors lead to deviations in the evaluation of treatment responses by the RECIST criteria, which rely on changes in tumor size on imaging. Therefore, exploring a more accurate method for efficacy assessment and prognostication of PDAC after NACT is of great clinical significance.

¹⁸F-FDG PET/CT integrates the dual advantages of functional metabolic and anatomical morphological imaging for tumors, and its quantitative analysis of metabolic parameters has demonstrated unique value in the diagnosis, staging, and monitoring of distant metastasis in PDAC[11–13]. Moreover, the PET Response Criteria in Solid Tumors version 1.0 (PERCIST 1.0), derived from this technique, has achieved significant progress in prognostic prediction for malignant tumors such as non-small cell lung cancer and lymphoma[14, 15]. Thus, this study aimed to investigate the prognostic value of ¹⁸F-FDG PET/CT metabolic parameters in PDAC patients receiving NACT, in order to provide a reliable imaging reference for the formulation of individualized clinical treatment strategies.

## 2. Materials and Methods

### 2.1 Study Population

We retrospectively collected the general data, clinicopathological data, and imaging data of pathologically confirmed PDAC patients in our hospital from December 2017 to December 2022. All patients received chemotherapy regimens recommended by the National Comprehensive Cancer Network (NCCN) and the College of American Pathologists.

The inclusion criteria were (1) pathologically confirmed PDAC by histopathology; (2) treatment-naive patients who did not receive anti-tumor therapy within 6 months before enrollment; (3) patients treated with NACT regimens for PDAC as recommended by the NCCN guidelines; and (4) patients who underwent ¹⁸F-FDG PET/CT, CT, and MRI scans within 2 weeks before and after NACT. The exclusion criteria were (1) missing original PET/CT, CT, or MRI data; (2) patients who received radiotherapy, immunotherapy, targeted therapy, or clinical trial regimens during NACT; (3) patients with other primary malignant tumors; and (4) patients who refused treatment or lost follow-up due to treatment refusal.

### 2.2 Study Design

All patients completed relevant examinations including MRI, contrast-enhanced CT, and serological tests upon admission. In accordance with the NCCN guidelines [1], all patients received more than 3 cycles of NACT, and completed ¹⁸F-FDG PET/CT, contrast-enhanced CT, and contrast-enhanced MR examinations within 2 weeks before and after standardized chemotherapy. The standardized medication regimens were as follows:

For the AG regimen (Gemcitabine plus nab-paclitaxel), nab-paclitaxel (125 mg/m^2^) was intravenously infused on days 1 and 8; Gemcitabine (1000 mg/m^2^) was intravenously infused on days 1 and 8, repeated every 3 weeks. For the FOLFIRINOX regimen, Oxaliplatin (85 mg/m^2^) was intravenously infused on day 1; Irinotecan (180 mg/m^2^) was intravenously infused on day 1; Leucovorin (400 mg/m^2^) was intravenously infused on day 1; 5-fluorouracil (400 mg/m^2^) was rapidly injected intravenously on day 1, followed by a continuous infusion of 5-fluorouracil (2400 mg/m^2^) for 46 hours, repeated every 2 weeks[16].

### 2.3 ¹⁸F-FDG PET/CT Scanning Protocol and Image Processing

Examinations were performed using a Siemens Biograph 64 PET/CT scanner (Siemens Healthineers, Erlangen, Germany), and image acquisition was performed strictly in accordance with the European Association of Nuclear Medicine ¹⁸F-FDG PET/CT tumor imaging procedure guidelines[17]. The ¹⁸F-FDG reagent had a radiochemical purity of more than 95%. Patients were instructed to fast for 4–6 hours before the examination, and after confirming a blood glucose level < 9.1 mmol/L, ¹⁸F-FDG was intravenously injected at a dose of 5.55 MBq/kg body weight. After injection, patients rested in a dark room for 50–60 minutes before whole-body ¹⁸F-FDG PET/CT scanning. Prior to scanning, patients were instructed to drink 800 mL of water to promote gastric filling and to void the bladder before imaging. The scanning range was set from the base of the skull to the midpoint of the femur, and whole-body PET/CT image acquisition was completed using a spiral CT scanning mode with the following parameters: tube voltage 120 kV, tube current 170 mA, slice interval 0.75 mm, and slice thickness 0.5 mm. PET images were iteratively reconstructed using the three-dimensional ordered subset expectation maximization method, and attenuation correction of PET images was completed by combining relevant information from CT scans.

The acquired PET/CT images were imported into LIFEx software (version 4.00) in Digital Imaging and Communications in Medicine (DICOM) format. The region of interest of the pancreatic target lesion was delineated with a threshold of 40% of the maximum standardized uptake value (SUVmax), and the software automatically measured tumor metabolism-related parameters including SUV, metabolic tumor volume (MTV), and total lesion glycolysis (TLG). All PET/CT image data were independently interpreted by two nuclear medicine physicians with at least 3 years of PET/CT diagnostic experience. In cases of discrepancies between the two physicians’ judgments, a departmental collective discussion was held to resolve the discrepancies. Meanwhile, the two physicians delineated the region of interest in combination with the lesion contours, and the final treatment response was determined by consensus after joint discussion, in accordance with the PERCIST 1.0 criteria[18].

The quantitative analysis results from the baseline PET/CT examination (before NACT) are referred to herein as PET1 variables (SUVmax_1_, MTV_1_, TLG_1_, standardized uptake value normalized to lean body mass [SUL]), and those from the second examination (after NACT) are referred to as PET2 variables (SUVmax_2_, MTV_2_, TLG_2_, SUL). The formula for calculating the percentage change of each parameter was as follows: ΔSUVmax% = ([SUVmax_1_–SUVmax_2_]/SUVmax_1)_ × 100%. The percentage changes of MTV, TLG, and SUL (ΔMTV%, ΔTLG%, and ΔSUL%, respectively) were calculated in the same manner.

According to the RECIST 1.1 criteria, patients with a complete response (CR) or partial response were classified into the radiological response group, and those with progressive disease or stable disease were classified into the radiological non-response group. Based on the 30% cut-off value of PERCIST 1.0, patients were categorized into a metabolic response group (complete or partial metabolic response) or a metabolic non-response group (progressive or stable metabolic disease).

### 2.4 Follow-up

The date of the first ¹⁸F-FDG PET/CT examination before NACT was set as the baseline. All patients were followed up for 3 years, with the last follow-up on October 1, 2025. Follow-up methods mainly included outpatient, telephone, and inpatient follow-up, with a frequency of once a month within 3 months after NACT completion; once every 3 months from 3 to 12 months after completion; and once every 6 months thereafter. Follow-up end points were defined as follows. Overall survival (OS) was calculated from the time from baseline to patient death or the last follow-up (in months). Progression-free survival (PFS) was calculated from the time from baseline to tumor recurrence, metastasis, or the last follow-up (in months)[19]. Loss to follow-up was defined as patients who failed to participate in two consecutive follow-ups and could not be contacted by telephone or messaging or patients who voluntarily abandoned follow-up. For patients who were lost to follow-up, the last follow-up date was used as the censoring time for survival analysis. A total of 44 patients were included in this study, with no loss to follow-up during the follow-up period. The follow-up duration ranged from 4 to 57 months, with a median duration of 23.5 months (interquartile range [IQR]: 15.0–31.0).

### 2.5 Statistical Analysis

SPSS version 27.0 (IBM Corp., Armonk, NY, USA) and R version 4.2.3 were used for statistical analysis . Categorical data are presented as numbers and percentages. For continuous data with a normal distribution, the mean and standard deviation are presented; otherwise, the median and IQR are shown. Intergroup comparisons of categorical data were performed using the chi-squared test or Fisher’s exact test. Time-dependent receiver operating characteristic curves were applied to calculate the optimal cut-off values of PET/CT metabolic parameters, and continuous data were converted into categorical data based on the cut-off values. The Kaplan–Meier method was used to plot survival curves, and the log-rank test was used to analyze intergroup differences. Univariate and multivariate Cox proportional hazards regression models were used to analyze prognostic factors, and variables with statistical significance in the univariate analysis were included in the multivariate analysis to identify independent factors affecting long-term survival and disease progression in patients. Unless otherwise specified, two-sided P-values < 0.05 were considered statistically significant.

## 3. Results

### 3.1 Baseline Characteristics and Overall Survival

A total of 163 patients were initially screened, and 44 eligible patients with complete follow-up were finally included after applying the inclusion and exclusion criteria. Their baseline characteristics are shown in Table 1. The median OS of the 44 patients was 21.0 months (range: 4–57 months), and the median PFS was 10.0 months (range: 2–36 months). At the end of follow-up, 29 patients (65.90%) had died, all patients (100.00%) developed disease progression, and the 3-year survival rate of the cohort was 22.72%.

**Table 1.** Baseline Clinical Characteristics of the Study Cohort (N=44)

| Variable | Characteristic |
| --- | --- |
| Sex (Male/Female) | 26/18 |
| Age (years) | 64.23±8.19 |
| Tumor Location (pancreatic head/body/tail) | 21/19/4 |
| Clinical Stage (Stage I/II/III + IV) | 3/6/35 |
| Surgical Resectability (BRPC/LAPC/UPC/RPC) | 16/23/5 |
| NACT Regimen (FOLFIRINOX/AG) | 16/28 |
| Pre-NACT CA19-9 (U/mL) | 260.30 (IQR, 65.35–616.50) |
| Post-NACT CA19-9 (U/mL) | 55.45 (IQR, 13.55–141.20) |
| Percentage Change* (%) | 66.00 (IQR, 36.00–91.00) |
| Pre-NACT CEA (U/mL) | 3.05 (IQR, 2.20–5.10) |
| Post-NACT CEA (U/mL) | 3.35 (IQR, 2.20–4.25) |
| Percentage Change* (%) | 5.02 (IQR, –50.14–32.00) |
| PERCIST 1.0 Classification<br>(*CMR+PMR/PMD+SMD*) | 29/15 |
| RECIST 1.1 Classification (*CR+PR/PD+SD*) | 22/22 |
| Subsequent Treatment (Radical resection/Other) | 24/20 |

AG, Gemcitabine plus nab-paclitaxel; BRPC, borderline resectable pancreatic cancer; CA19-9, carbohydrate antigen 19-9; CEA, carcinoembryonic antigen; CMR, complete metabolic response; CR, complete response; FOLFIRINOX, fluorouracil, leucovorin, irinotecan, and oxaliplatin; IQR, interquartile range; LAPC, locally advanced pancreatic cancer; NACT, neoadjuvant chemotherapy; PDAC, pancreatic ductal adenocarcinoma; PD, progressive disease; PMD, progressive metabolic disease; PMR, partial metabolic response; PERCIST, PET Response Criteria in Solid Tumors; PR, partial response; RECIST, Response Evaluation Criteria in Solid Tumors; RPC, resectable pancreatic cancer; SD, stable disease; SMD, stable metabolic disease; UPC, unresectable pancreatic cancer

### 3.2 Comparative Prognostic Value of PERCIST 1.0 and RECIST 1.1 for Long-term Outcome

Kaplan–Meier survival analysis showed that patients with a metabolic response according to PERCIST 1.0 had a significantly better prognosis than those with a metabolic non-response (3-year survival rate: 31.03% vs. 0.00%; log-rank *P* = 0.018; Figure 1). The chi-squared test confirmed that there was a statistically significant difference in the 3-year survival rate between the PERCIST 1.0 subgroups (*χ^2^* = 5.852, *P* = 0.018), while no significant difference was observed between the RECIST 1.1 subgroups (Table 2).

**Fig 1.**
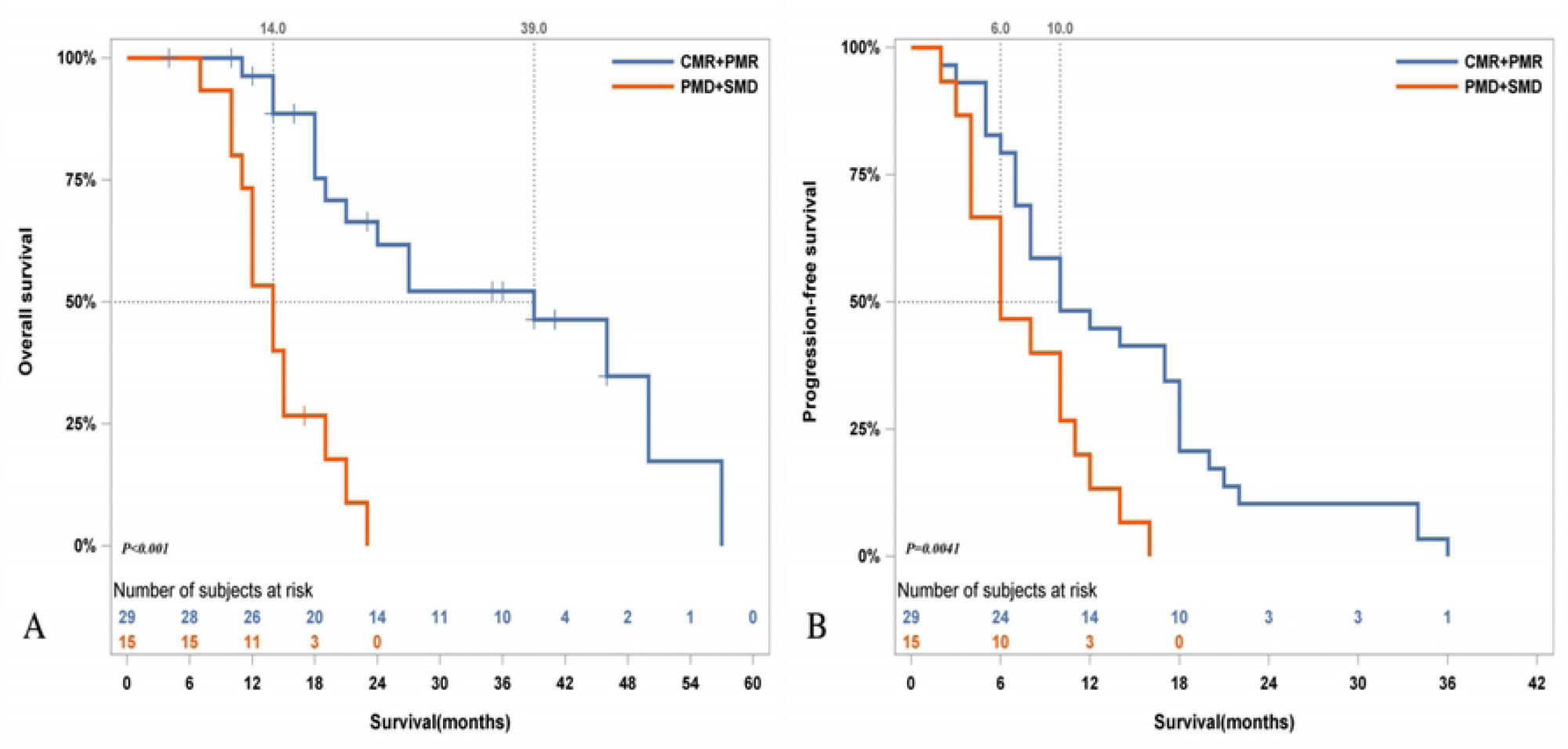
Kaplan–Meier Survival Curves for OS and PFS in Patients with PDAC Stratified by PERCIST 1.0 Criteria. (A) The median OS of the metabolic response group according to PERCIST 1.0 was significantly higher than that of the metabolic non-response group (39.0 months vs. 14.0 months, *P* < 0.001); (B) The median PFS of the metabolic response group according to PERCIST 1.0 was higher than that of the metabolic non-response group (10.0 months vs. 6.0 months, *P* = 0.004). Abbreviations: OS, Overall survival; PFS, Progression-free survival; PDAC, Pancreatic ductal adenocarcinoma; PERCIST 1.0, PET Response Criteria in Solid Tumors version 1.0

**Table 2.** Predictive Analysis of 3-Year Survival Rates in Patients with PDAC Receiving NACT.

| Evaluation | Number | >36 | months | $\leq 36$ | months | $\chi^2$ | $P$ |
| --- | --- | --- | --- | --- | --- | --- | --- |
| Criterion | of Cases | (n=10) |  | (n=34) |  |  |  |
| PERCIST 1.0 |  |  |  |  |  |  |  |
| Metabolic response group | 29 | 9 (31.03) | | 20 (68.97) | | $\chi^2=5.852$ | 0.018 |
| Metabolic non-response group | 15 | 0 (0.00) |  | 15 (100.00) |  |  |  |
| RECIST 1.1 |  |  |  |  |  |  |  |
| Radiological response group | 22 | 5 (22.73) | | 17 (77.27) | | $\chi^2=0.049$ | 1.000 |
| Radiological non-response group | 22 | 4 (18.18) |  | 18 (81.82) |  |  |  |

NACT, neoadjuvant chemotherapy; PDAC, pancreatic ductal adenocarcinoma; PERCIST, PET Response Criteria in Solid Tumors; RECIST, Response Evaluation Criteria in Solid Tumors

Univariate Cox regression analysis showed that the PERCIST 1.0 results were significantly correlated with OS and PFS (*P* < 0.001, *P* = 0.008), while the RECIST 1.1 results had no correlation with prognosis (*P* = 0.750, *P* = 0.414; Table 3).

**Table 3.**
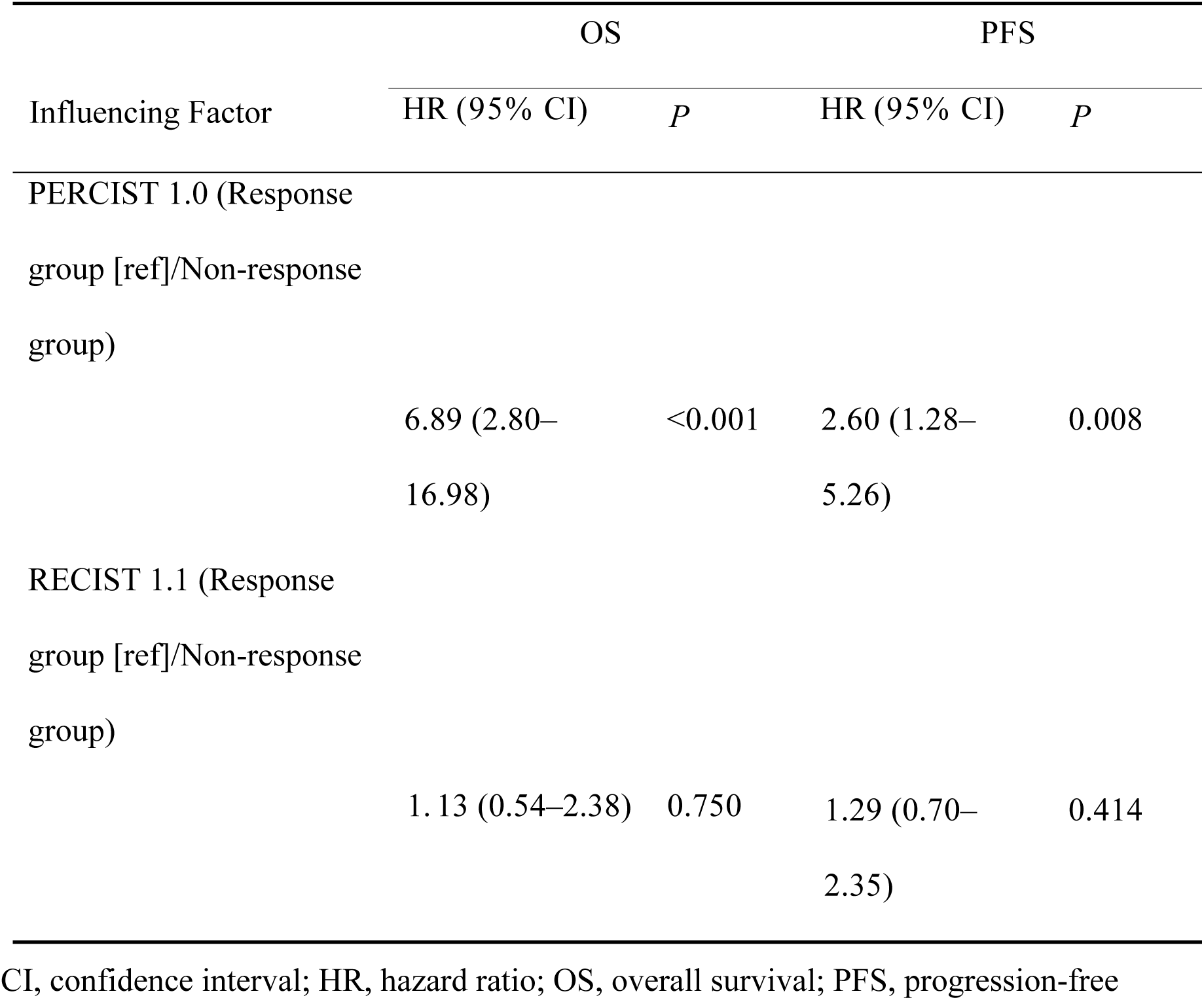
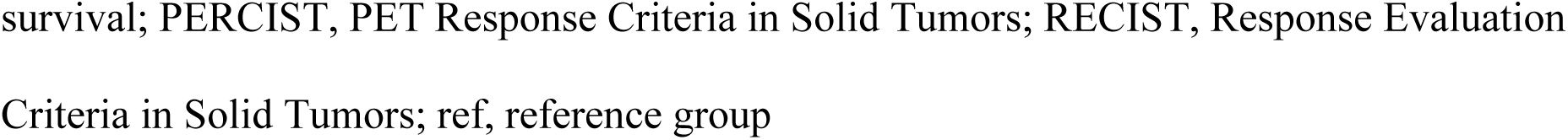
Univariate Cox Regression Analysis of Prognostic Factors.

| Influencing Factor | OS |  | PFS |  |
| --- | --- | --- | --- | --- |
|  | HR (95% CI) | *P* | HR (95% CI) | *P* |
| PERCIST 1.0 (Response group [ref]/Non-response group) | 6.89 (2.80–16.98) | <0.001 | 2.60 (1.28–5.26) | 0.008 |
| RECIST 1.1 (Response group [ref]/Non-response group) | 1.13 (0.54–2.38) | 0.750 | 1.29 (0.70–2.35) | 0.414 |

CI, confidence interval; HR, hazard ratio; OS, overall survival; PFS, progression-free
survival; PERCIST, PET Response Criteria in Solid Tumors; RECIST, Response Evaluation Criteria in Solid Tumors; ref, reference group

Multivariate Cox regression analysis revealed that a metabolic non-response after NACT was an independent risk factor for poor prognosis; compared with metabolic responders, patients classified with either stable metabolic disease or progressive metabolic disease using the PERCIST 1.0 criteria had shorter OS and a higher risk of disease progression (hazard ratio [HR] = 6.23, 95% confidence interval [CI]: 2.12–18.29, *P* < 0.001; HR = 4.30, 95% CI: 1.63– 11.35, *P* = 0.003), as detailed in Table 3. The survival analysis results of the metabolic response and non-response groups are shown in Figure 1.

### 3.3 Prognostic Value of ¹⁸F-FDG PET/CT Metabolic Parameters

Time-dependent receiver operating characteristic curve analysis was first used to determine the optimal cut-off values of each ¹⁸F-FDG PET/CT metabolic parameter (Table 4). Among these parameters, SUL-related parameters (SULpeak_2_, ΔSULpeak%) had the highest concordance index, with 0.686 (95% *CI*: 0.655–0.717) and 0.738 (95% *CI*: 0.712–0.764), respectively, which were significantly superior to traditional metabolic parameters such as SUV, MTV, and TLG (all concordance index values < 0.7).

**Table 4.**
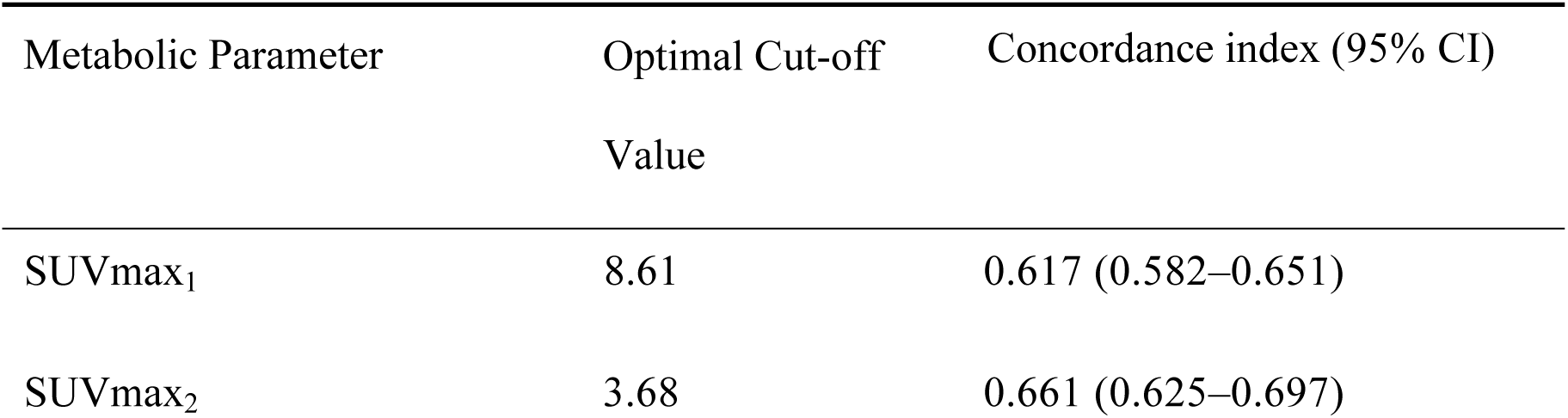
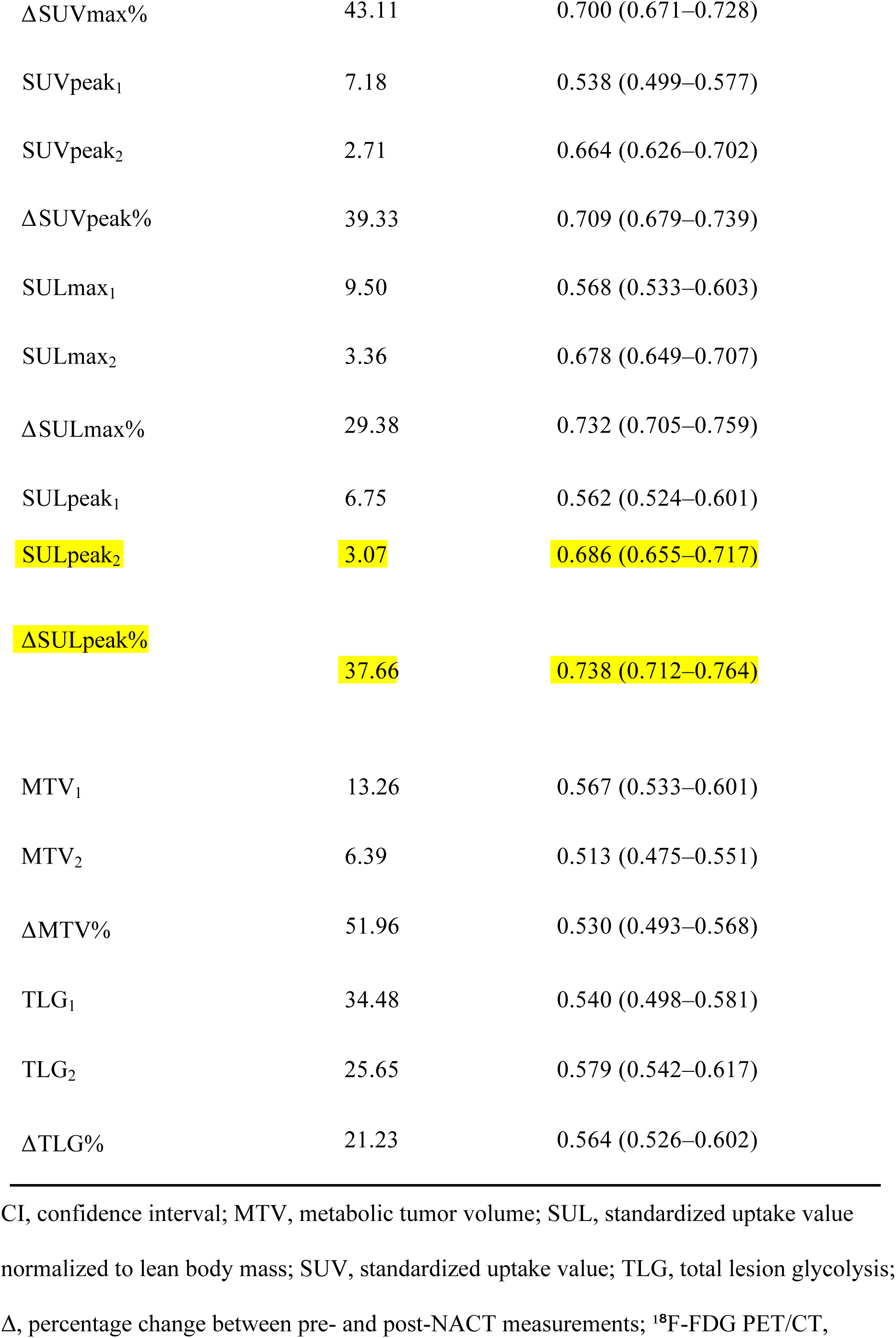
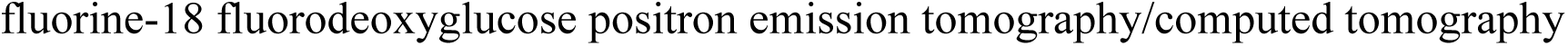
Optimal Cut-off Values of ¹⁸F-FDG PET/CT Metabolic Parameters.

| Metabolic Parameter | Optimal Cut-off Value | Concordance index (95% CI) |
| --- | --- | --- |
| SUVmax<sub>1</sub> | 8.61 | 0.617 (0.582–0.651) |
| SUVmax<sub>2</sub> | 3.68 | 0.661 (0.625–0.697) |
| ΔSUV<sub>max</sub>% | 43.11 | 0.700 (0.671–0.728) |
| SUV<sub>peak1</sub> | 7.18 | 0.538 (0.499–0.577) |
| SUV<sub>peak2</sub> | 2.71 | 0.664 (0.626–0.702) |
| ΔSUV<sub>peak</sub>% | 39.33 | 0.709 (0.679–0.739) |
| SUL<sub>max1</sub> | 9.50 | 0.568 (0.533–0.603) |
| SUL<sub>max2</sub> | 3.36 | 0.678 (0.649–0.707) |
| ΔSUL<sub>max</sub>% | 29.38 | 0.732 (0.705–0.759) |
| SUL<sub>peak1</sub> | 6.75 | 0.562 (0.524–0.601) |
| SUL<sub>peak2</sub> | 3.07 | 0.686 (0.655–0.717) |
| ΔSUL<sub>peak</sub>% | 37.66 | 0.738 (0.712–0.764) |
| MTV<sub>1</sub> | 13.26 | 0.567 (0.533–0.601) |
| MTV<sub>2</sub> | 6.39 | 0.513 (0.475–0.551) |
| ΔMTV% | 51.96 | 0.530 (0.493–0.568) |
| TLG<sub>1</sub> | 34.48 | 0.540 (0.498–0.581) |
| TLG<sub>2</sub> | 25.65 | 0.579 (0.542–0.617) |
| ΔTLG% | 21.23 | 0.564 (0.526–0.602) |

CI, confidence interval; MTV, metabolic tumor volume; SUL, standardized uptake value normalized to lean body mass; SUV, standardized uptake value; TLG, total lesion glycolysis; $\Delta$ , percentage change between pre- and post-NACT measurements; <sup>18</sup>F-FDG PET/CT,
fluorine-18 fluorodeoxyglucose positron emission tomography/computed tomography

Univariate Cox regression analysis showed that patients with a post-NACT *SUL*peak_2_ > 3.07 had significantly shorter OS and PFS (median OS: 15.0 months vs. 27.0 months, *P* = 0.007; median PFS: 6.0 months vs. 16.0 months, *P* < 0.001); and patients with a ΔSULpeak% > 37.66% had a significantly lower risk of death and disease progression (HR = 0.24, 95% CI: 0.11–0.53, *P* < 0.001; HR = 0.43, 95% CI: 0.22–0.84, *P* = 0.013). Among other parameters, only ΔTLG% > 21.23% was associated with longer OS (median OS: 27.0 months vs.14.0 months, *P* = 0.031), but it had no predictive value for PFS (*P* = 0.713), as detailed in Table 5.

**Table 5.**
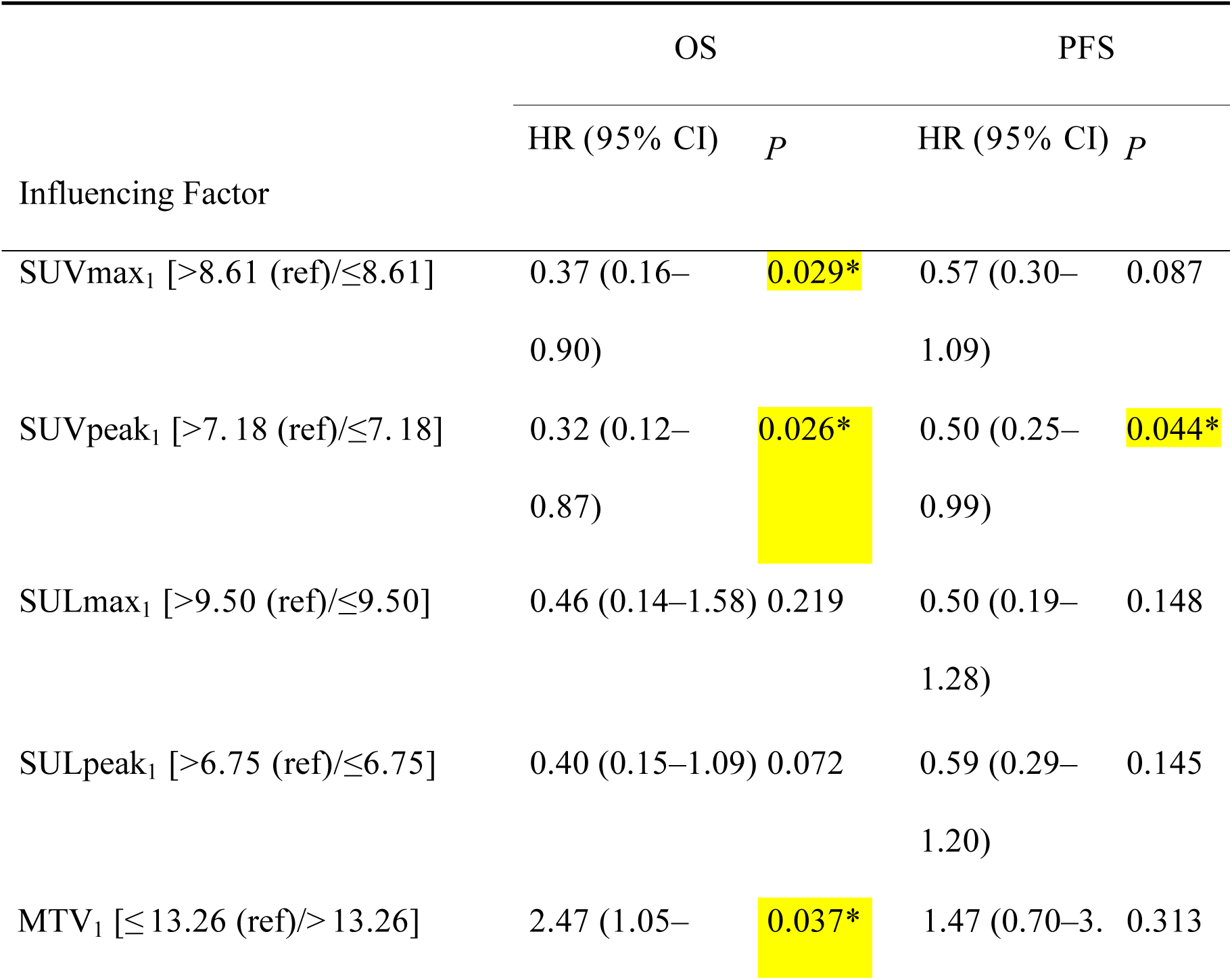
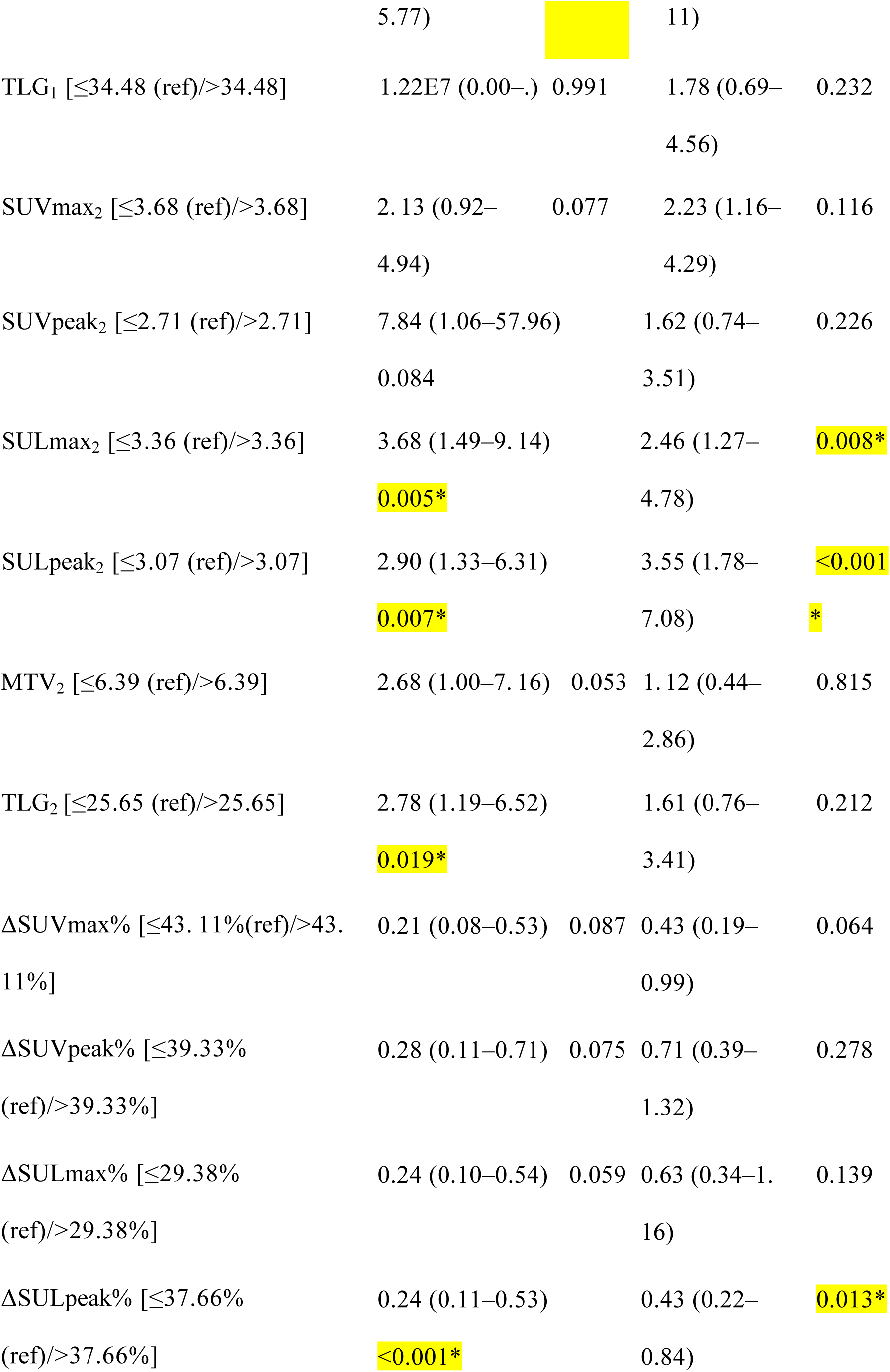
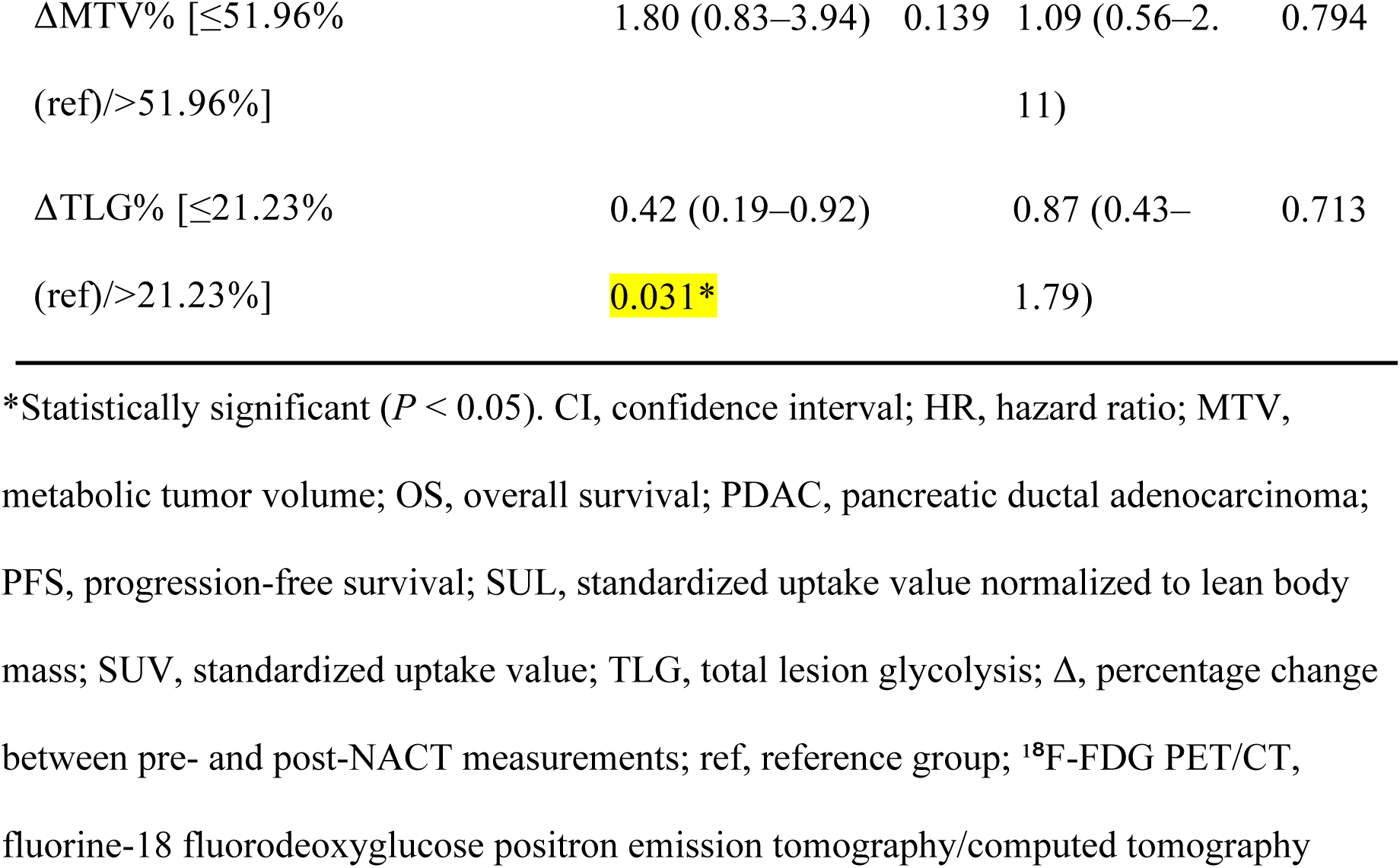
Univariate Cox Regression Analysis of ¹⁸F-FDG PET/CT Metabolic Parameters for Prognosis in PDAC.

Further multivariate Cox regression analysis confirmed that values of *SUL*peak_2_ > 3.07 and ΔSULpeak% ≤ 37.66% were independent risk factors for poor prognosis (Table 6). Kaplan–Meier survival curves further verified the above results (Figure 2).

**Fig 2.**
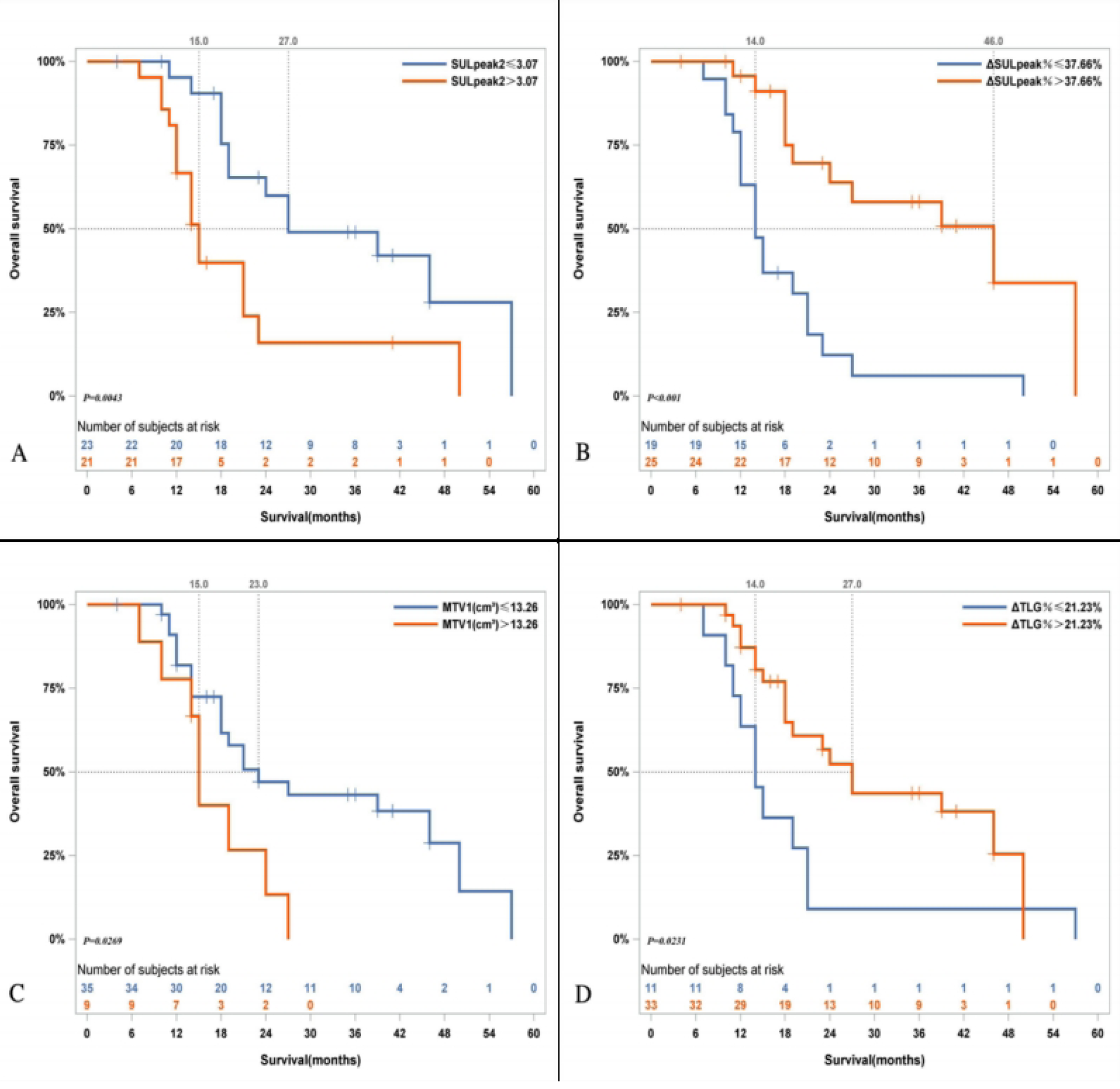
Kaplan–Meier Survival Curves for OS and PFS in Patients with PDAC Stratified by Key ¹⁸F-FDG PET/CT Metabolic Parameters. (A) A post-NACT SULpeak > 3.07 was associated with lower median OS than an SULpeak ≤ 3.07 (15.0 months vs. 27.0 months, *P* = 0.004). (B) A ΔSULpeak% > 37.66% was associated with a significantly higher median OS than a ΔSULpeak% ≤ 37.66% (46.0 months vs. 14.0 months, *P* < 0.001). (C) A pre-NACT MTV > 13.26 cm^3^ was associated with a lower median OS than an MTV ≤ 13.26 cm^3^ (15.0 months vs. 23.0 months, *P* = 0.027). (D) A ΔTLG% > 21.23% was associated with a higher median OS than a ΔTLG% ≤ 21.23% (27.0 months vs. 14.0 months, *P* = 0.023). (E) A post-NACT SULpeak > 3.07 was associated with a significantly lower median PFS than an SULpeak ≤ 3.07 (6.0 months vs. 16.0 months, *P* < 0.001). (F) A ΔSULpeak% > 37.66% was associated with a higher median PFS than a ΔSULpeak% ≤ 37.66% (12.0 months vs. 8.0 months, *P* = 0.008).

**Table 6.**
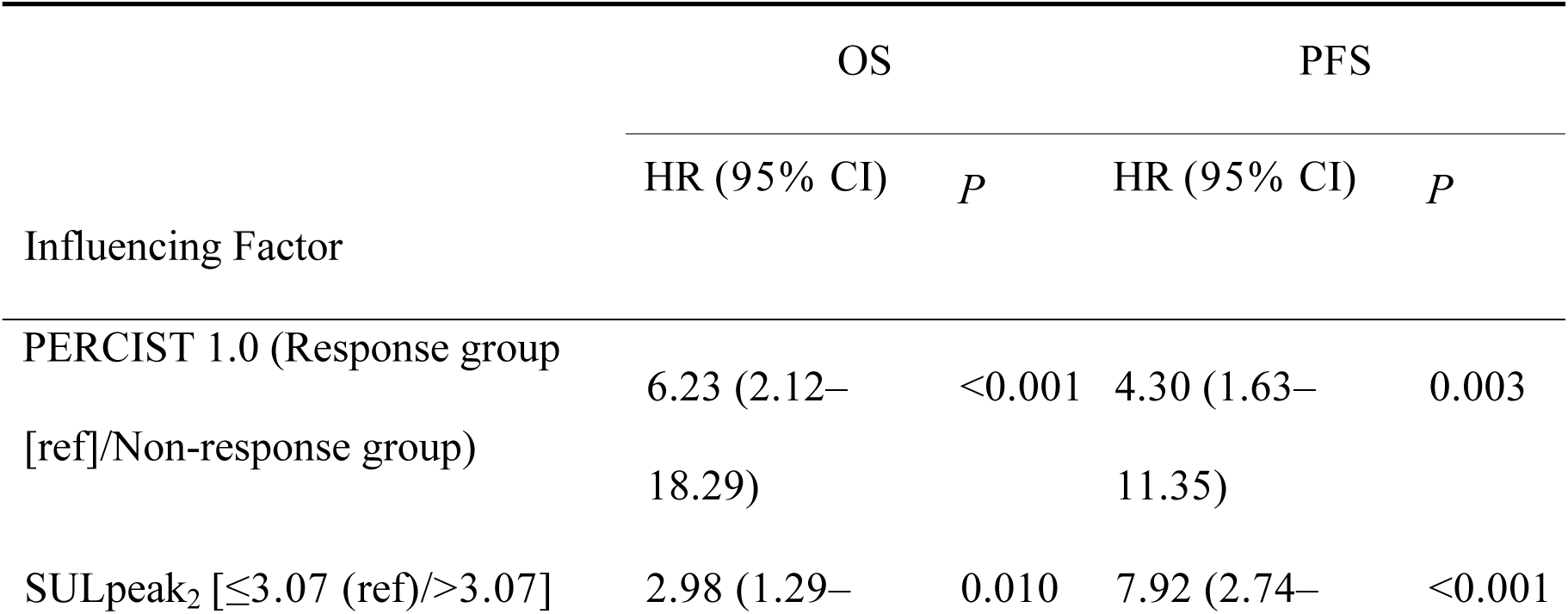
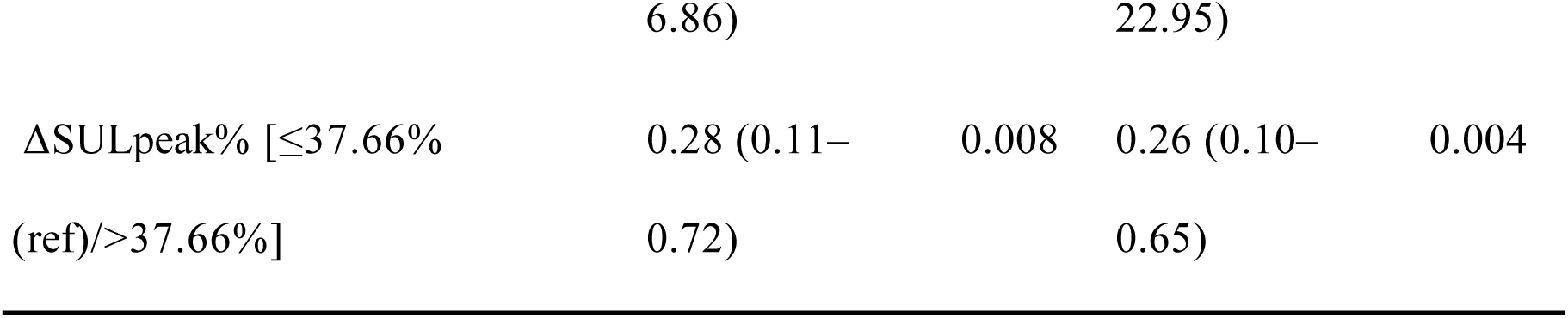
Multivariate Cox Regression Analysis of Factors Affecting OS and PFS in Patients with PDAC.

## 4. Discussion

This study retrospectively analyzed clinical and imaging data of 44 patients with PDAC receiving NACT to evaluate the prognostic value of PERCIST 1.0 and quantitative metabolic parameters derived from ¹⁸F-FDG PET/CT imaging data. The results confirmed that PERCIST 1.0 could more accurately assess treatment responses and predict long-term survival than RECIST 1.1, and it identified SULpeak_2_ and ΔSULpeak% as the core metabolic indicators for evaluating prognosis in this patient population. These findings provide an important functional imaging basis for addressing the limitations of morphological criteria in assessing treatment efficacy in PDAC and offer quantifiable reference indicators for individualized prognostic stratification and clinical interventions.

PERCIST 1.0 has demonstrated its value in the evaluation of treatment efficacy and prognosis for many types of solid malignant tumors[20–22]. In this study, the 3-year survival rate of patients in the PERCIST 1.0 metabolic response group reached 31.03%, which was substantially higher than that in the non-response group, and multivariate Cox regression analysis confirmed that a metabolic non-response was an independent risk factor for both OS (HR = 6.23) and PFS (HR = 4.30). Recently, Jiang[23] et al. reported similar findings in neuroendocrine tumors, where PERCIST 1.0 identified higher objective response and disease control rates than morphological criteria. Our survival analysis showed that treatment responses according to the RECIST 1.1 criteria were not significantly associated with 3-year survival rates, OS, or PFS, which is consistent with previous research[24, 25]. Chatterjee[26] et al. suggested that this is due to significant fibrosis in patients with PDAC receiving NACT, which renders morphological assessment unable to reflect the true biological behavior of PDAC post-chemotherapy.

This study found that ¹⁸F-FDG PET/CT metabolic parameters possess significant prognostic value in patients with PDAC receiving NACT; SUL-related parameters demonstrated significantly better predictive efficacy than traditional parameters such as SUV, MTV, and TLG, among which a post-NACT SULpeak > 3.07 and a ΔSULpeak% ≤ 37.66% were independent predictors of poor prognosis. SUL has shown prominent advantages in recent studies. Hennessy[27] et al. confirmed that an SULmax ≤ 3 was associated with better survival outcomes in patients with breast cancer receiving NACT. Zhang[28] et al. also showed that a SULpeak reduction of more than 63.92% could serve as a reference value for identifying patients who might be candidates for non-operative management after NACT. Additionally, the present study found that ΔTLG% had only modest predictive value for OS and no significant association with PFS in PDAC, with inferior predictive performance compared to SULpeak. This may be because a higher SUL value indicates enhanced tumor metabolic activity and accelerated cell proliferation, which are strongly associated with increased tumor invasiveness and poor prognosis[29].

Regarding changes in tumor markers, carbohydrate antigen 19-9 (CA19-9) levels decreased significantly in patients with PDAC after NACT (median percentage change: 66.00%), whereas carcinoembryonic antigen levels showed no substantial change. Furthermore, the levels and percentage changes of CA19-9 and carcinoembryonic antigen before and after treatment were not associated with OS and PFS (Supplementary Table S1), which aligns with previous studies[30]. This suggests that re-evaluation with PET/CT after NACT is beneficial for patients with PDAC, particularly for those classified as having progressive disease by RECIST 1.1 despite stable or decreasing CA19-9 levels; in such cases, PERCIST 1.0 may represent a more valuable method for prognostic assessment.

In this study, the median OS of the 44 patients was only 21.0 months, with a 3-year survival rate of 22.72%, and all patients experienced disease progression by the end of follow-up. These results underscore that the overall prognosis for patients with locally advanced and advanced PDAC remains poor even after standardized NACT. The application of PERCIST 1.0 and SUL-related metabolic parameters can facilitate the early identification of patients who might benefit from chemotherapy and those at high risk of poor outcomes, providing a crucial basis for the timely adjustment of clinical treatment regimens. For patients exhibiting a metabolic non-response or meeting the criteria of a post-NACT SULpeak_2_ > 3.07 and a Δ SULpeak% ≤ 37.66%, early consideration of alternative chemotherapy regimens or combined comprehensive interventions, such as targeted therapy or immunotherapy, may be warranted to improve prognosis[31].

This study has certain limitations: first, it was a retrospective study with inherent selection bias; second, the sample size was small and the study lacked external validation with multicenter data; third, the latest research related to PET/MRI was not incorporated or cited. Future studies should conduct large-sample, multicenter cohort investigations to further validate and refine the findings of this study.

## 5. Conclusions

This study confirmed that PERCIST 1.0, based on ¹⁸F-FDG PET/CT data, demonstrated superior prognostic stratification than RECIST 1.1 for patients with PDAC after NACT and that the evaluation results using RECIST 1.1 showed no correlation with prognosis. Quantitative ¹⁸F-FDG PET/CT metabolic parameters showed significant prognostic value, and a post-NACT SULpeak > 3.07 and a ΔSULpeak% ≤ 37.66% were shown to be independent predictive factors for poor prognosis in these patients.

## Declarations

### Ethics approval and consent to participate

This research was conducted in compliance with the principles outlined in the Declaration of Helsinki and data combined with clinical patient information received approval from the ethics committee of Zhejiang Provincial People’s Hospital (Approval NO: QT2026046). Due to the anonymity of patient data, the need for informed consent was waived.

### Human Ethics

The study strictly adhered to the ethical principles of the Declaration of Helsinki, and all experimental protocols were consistent with the approved ethical review opinion.

### Consent for publication

All participants provided written consent for the publication of their data.

### Availability of supporting data

Data can be acquired from the corresponding author on reasonable request.

### Authors’ contributions

Linghe Zhang and Luqiang Jin conceived and designed the study; collected data; analyzed results; wrote and revised the manuscript; and approved the final version.

## Acknowledgments

We would like to thank and express our gratitude to Editage for editorial assistance.

## Supporting Information Captions

**Supplementary Table S1. Univariate Cox Regression Analysis of Serum Tumor Markers for Predicting Prognosis in Patients with Pancreatic Cancer**

